# Identification of Hypertrophic Cardiomyopathy on Electrocardiographic Images with Deep Learning

**DOI:** 10.1101/2023.12.23.23300490

**Authors:** Veer Sangha, Lovedeep Singh Dhingra, Arya Aminorroaya, Philip M Croon, Nikhil V Sikand, Sounok Sen, Matthew W Martinez, Martin S Maron, Harlan M Krumholz, Folkert W Asselbergs, Evangelos K Oikonomou, Rohan Khera

## Abstract

Hypertrophic cardiomyopathy (HCM) is frequently underdiagnosed. While deep learning (DL) models using raw electrocardiographic (ECG) voltage data can enhance detection, their use at the point-of-care is limited. Here we report the development and validation of a DL model that detects HCM from images of 12-lead ECGs across layouts. The model was developed using 124,553 ECGs from 66,987 individuals at the Yale New Haven Hospital (YNHH), with HCM features determined by concurrent imaging (cardiac magnetic resonance [CMR] or echocardiography). External validation included ECG images from MIMIC-IV, Amsterdam University Medical Center (AUMC), and UK Biobank, where HCM was defined by CMR (YNHH, MIMIC-IV, AUMC) and diagnosis codes (UK Biobank). The model demonstrated robust performance across image formats and sites (AUROCs: 0.95 internal testing; 0.94 MIMIC-IV; 0.92 AUMC; 0.91 UK Biobank). Discriminative features localized to anterior/lateral leads (V4-V5) regardless of layout. This approach enables scalable, image-based screening for HCM across clinical settings.

## INTRODUCTION

While hypertrophic cardiomyopathy (HCM) is among the leading causes of sudden cardiac death in young and middle-aged adults, scalable solutions for screening for the disease have remained elusive.^1,2^ HCM is a genetically determined disease that affects up to 1 in every 200 people globally.^3,4^ Additionally, HCM is an autosomal dominant disease expected to occur with equal frequency by gender, however women achieve clinical recognition less frequently than men. An early clinical diagnosis of HCM would potentially enable regular healthcare follow-up, rigorous cardiovascular risk management, and timely initiation of highly effective risk-reducing therapies for the population at large and potentially impact diagnostic disparities by gender.^2,5^ The diagnosis of HCM relies on cardiac imaging, such as echocardiography and cardiac magnetic resonance imaging (CMR).^5,6^ However, given the requirement of expensive technology and extensive clinical expertise in deploying and interpreting these modalities, using echocardiography or CMR as a screening strategy for earlier diagnosis is not feasible.^6,7^

With the inaccessibility of advanced cardiac imaging, artificial intelligence (AI)-enhanced interpretation of electrocardiograms (AI-ECG) has been proposed as an alternative for the early detection of HCM.^8,9^ While ECG abnormalities, such as prominent Q waves, repolarization changes, left axis deviation, or giant negative T waves, can be apparent in over 90% of patients with the disease, these changes are not specific to HCM.^10,11^ Deep learning-based approaches, such as convolutional neural networks (CNNs), can leverage HCM-specific pathological signatures to identify people with the disease using clinical ECGs.^8,9,12^ Current models, however, use raw ECG voltage data as the inputs, often stored in vendor-specific formats and rarely accessible to clinicians at the point of care. Moreover, case selection in some previous model development has not consistently followed clinical definitions of HCM, often identifying HCM with a combination of diagnosis codes and other administrative data elements, such as visits to the HCM clinic.^9,12^ Such approaches to phenotyping in the electronic health record are prone to misclassification and variability due to vast differences in health-system-specific coding practices for HCM.^13,14^ Therefore, there is an unmet need for the development of models that use ubiquitous and interoperable data formats for disease diagnosis and rely on objective imaging-based features for defining the presence of disease to enable AI-ECG’s use as a practical, generalizable, and scalable screening modality for HCM.

In this study, we report the development and validation of a deep learning-based approach for identifying CMR-confirmed HCM using images of 12-lead ECGs.

## RESULTS

### Study Population

We used a total of 126,203 12-lead ECGs from 68,109 patients at Yale New Haven Hospital (YNHH) collected between 2012 and 2021 for development and internal testing. The earliest magnetic resonance imaging (MRI) reports between 2013 and 2022 containing any mention of HCM were identified, and 779 patients were identified as having confirmed HCM, and 125 as possible HCM following review by four clinicians. The data from these patients were split into train, validation, and test sets at a patient level, as described in the methods. Briefly, only MRI-confirmed HCM cases were retained in validation and testing, while possible HCM cases based on MRI, and ECGs from patients with transthoracic echocardiograms (TTE) suggestive of severe ventricular hypertrophy (LVH) defined by interventricular septal thickness in diastole (IVSd) of greater than 15 mm were used to augment the robustness of HCM phenotype in training.

Individuals in the model development population (training and validation sets) had a median age of 63.2 years (IQR 51.2-74.1) at the time of ECG recording, and 33,257 (49.6%) were women. Overall, 48,475 (72.4%) were non-Hispanic White, 7978 (11.9%) were non-Hispanic Black, 6029 (9.0%) were Hispanic, 1300 (1.9%) were Asian, 376 (0.6%) were from other races, and information was missing for 2829 (4.2%). In the development population, there were 12,680 ECGs from 4745 patients with CMR-confirmation of HCM or echocardiographic parameters consistent with HCM **(Extended Table 1)**.

### Image-based Detection of HCM

We generated ECG images to mimic the variation seen in ECG layouts in the real-world. Our methodology, which mimics the processing steps ECG machines use to convert acquired waveform signals to printed outputs has been described previously.^15–19^ Briefly, given that real-world ECG images vary in appearance, each ECG in the training dataset was randomly plotted in one of several ECG formats, encapsulating different variations, including lead layout, color scheme, lead label font, size and position, and grid and signal line width (**Figure 1, Extended Figure 1**). In addition, images were randomly rotated between -10 and +10 degrees before input. The inclusion of several layouts, as well as image types was done to prevent overfitting to certain formats, or styles of ECG images, and to create a more versatile algorithm capable of accepting inputs from the many types of image capture used at the point of care. Further details are described in Methods, Image Generation. During evaluation, we assessed ECG images plotted in the four formats used in previous studies (**Extended Figure 2**).

**Figure 1.**
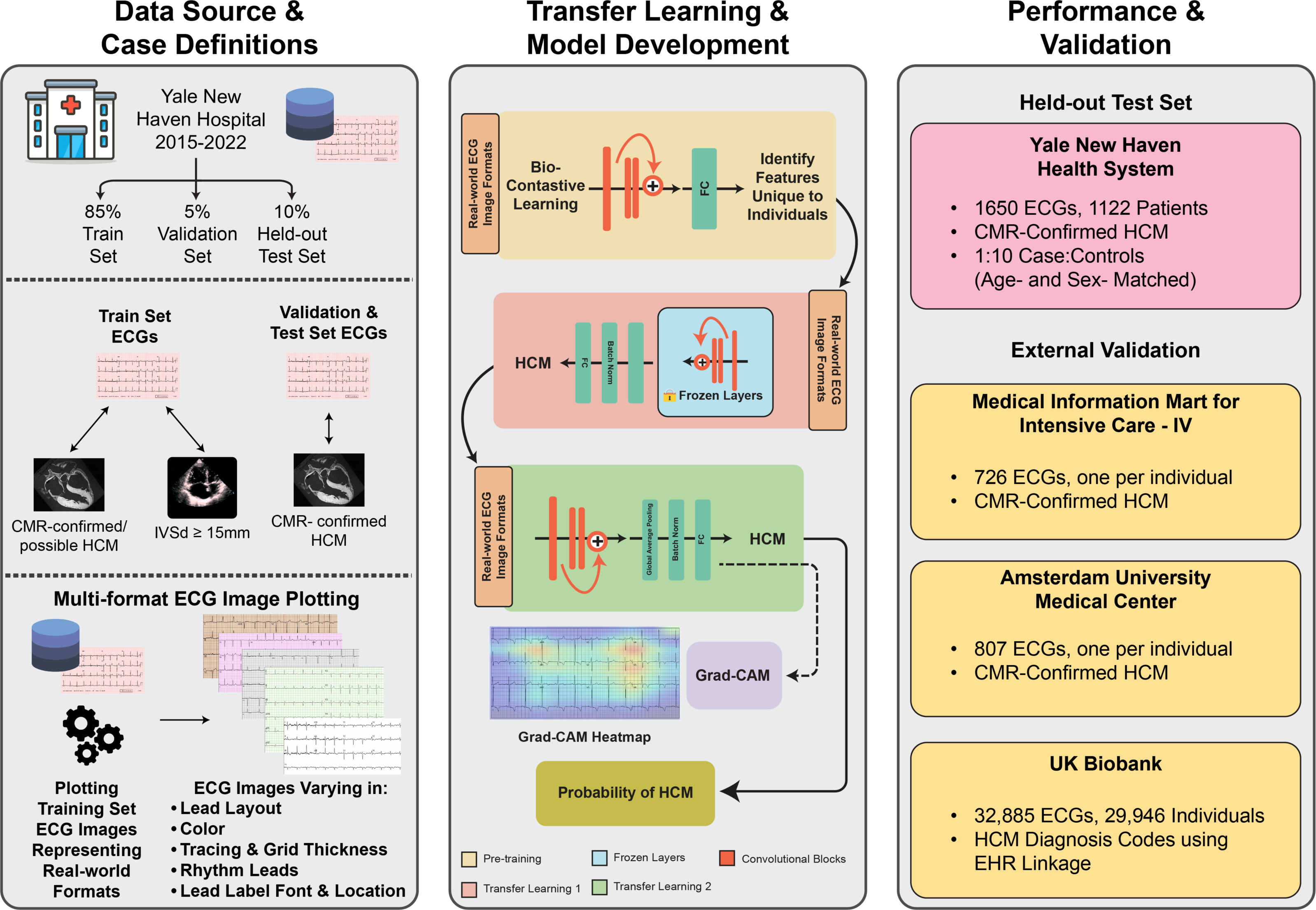
Model Development and Study Design. Abbreviations: CMR, Cardiac Magnetic Resonance Imaging; HCM, Hypertrophic Cardiomyopathy; IVSd, End-diastolic Interventricular Septal Thickness

**Figure 2.**
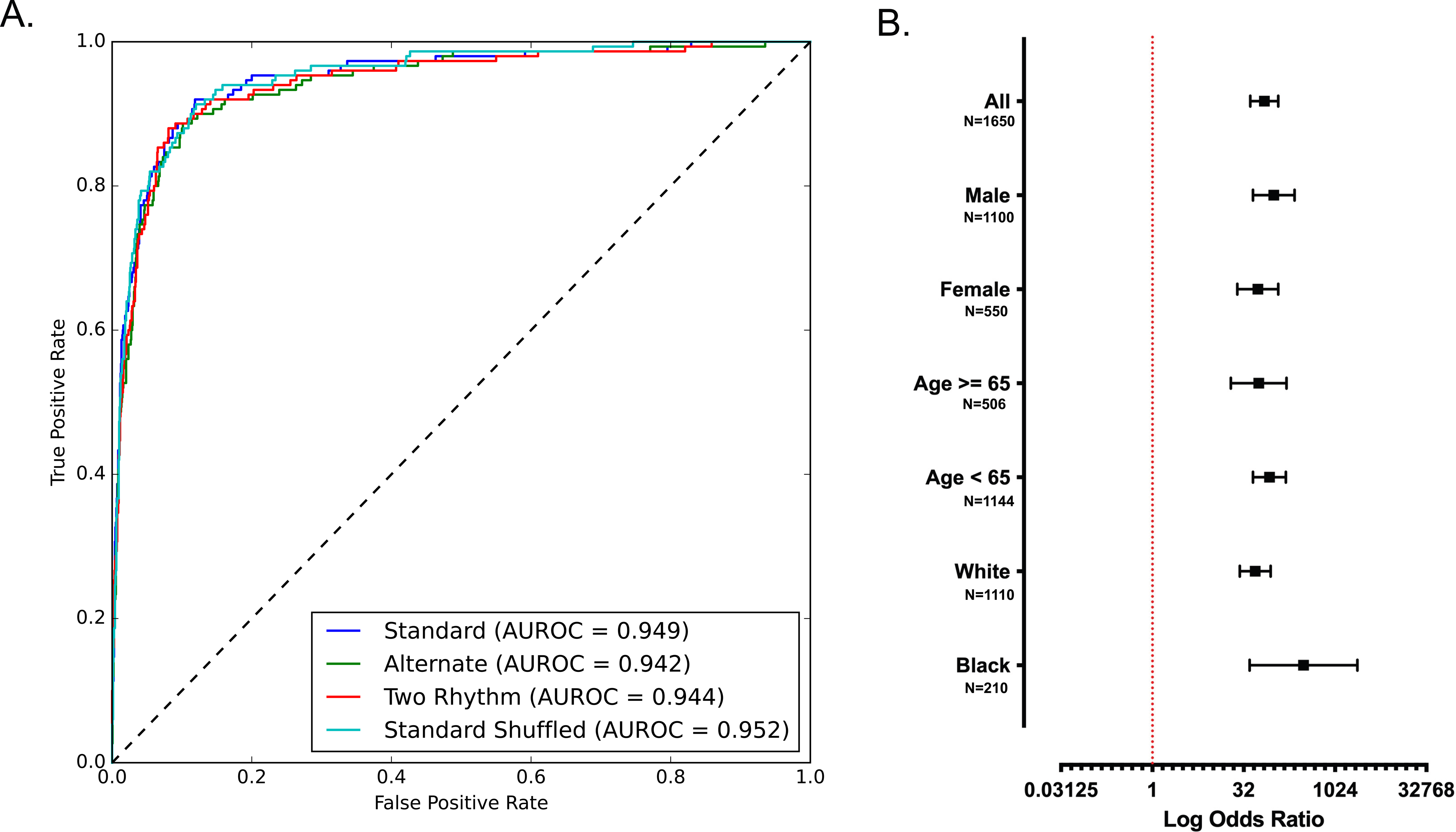
Model performance measures (A) Receiver operating characteristic curves across image formats in the held-out test set. B) Diagnostic odds ratios across age, gender, and race subgroups on standard format images in the held-out test set. Abbreviations: AUROC, area under receiver-operating characteristic curve. The box plots display the odds ratio, with the upper whiskers extending to 95^th^ percentile of the odds ratio, and the lower whiskers extending to the 5^th^ percentile of the odds ratio.* *Note: Diagnostic odds ratios were not available for Hispanic, Asian, or other Races because there were no FN screens in the held-out test set

To identify HCM signatures from these images, we trained a convolutional neural network model based on the EfficientNet-B3 architecture (see Methods, Model Architecture and Training for details).^20^ Before performing supervised fine tuning for detecting HCM, we initialized the model using weights from a self-supervised biocontrastive learning approach, in which the model was trained to identify individual patient-specific patterns.^21^ This allowed the model to learn features of HCM efficiently from the examples in the training cohort. Finally, to ensure that model learning was not affected by the lower frequency of HCM, we used a weighted binary cross-entropy loss function. Of note, the model can be publicly accessed at https://www.cards-lab.org/ecgvision-hcm for research use.

In the age- and sex-matched held-out test set comprising standard format images, the model for detecting HCM achieved an AUROC of 0.95 (**Figure 2**). A probability threshold for predicting HCM was chosen based on a sensitivity of 0.90 or higher in the validation subset.

With this threshold, the model had sensitivity and specificity of 0.89 and 0.90 in the held-out test set. Overall, an ECG suggestive of HCM indicated 70-fold higher odds (OR 69.9, 95% CI, 41.1 – 119.0) of HCM (**Figure 2**). The model’s performance was comparable across subgroups of age, sex, and race (**Extended Table 2, Table S1** and **Figure 2**). The model performance was also comparable across the four original layouts of ECG images in the held-out set with an AUROC of 0.94 – 0.95 for detecting HCM. Sensitivity analyses demonstrated consistent model performance on ECGs without paced rhythms, atrial fibrillation and flutter, conduction disorders, and in the presence of LVH (**Extended Table 2**). AI defined probabilities of HCM were longitudinally consistent amongst all 779 ECGs from the 59 patients in the held-out test set (**Extended Figure 3**), indicating that model identified changes in myocardial physiology associated with the HCM phenotype at several stages of disease progression.

**Figure 3.**
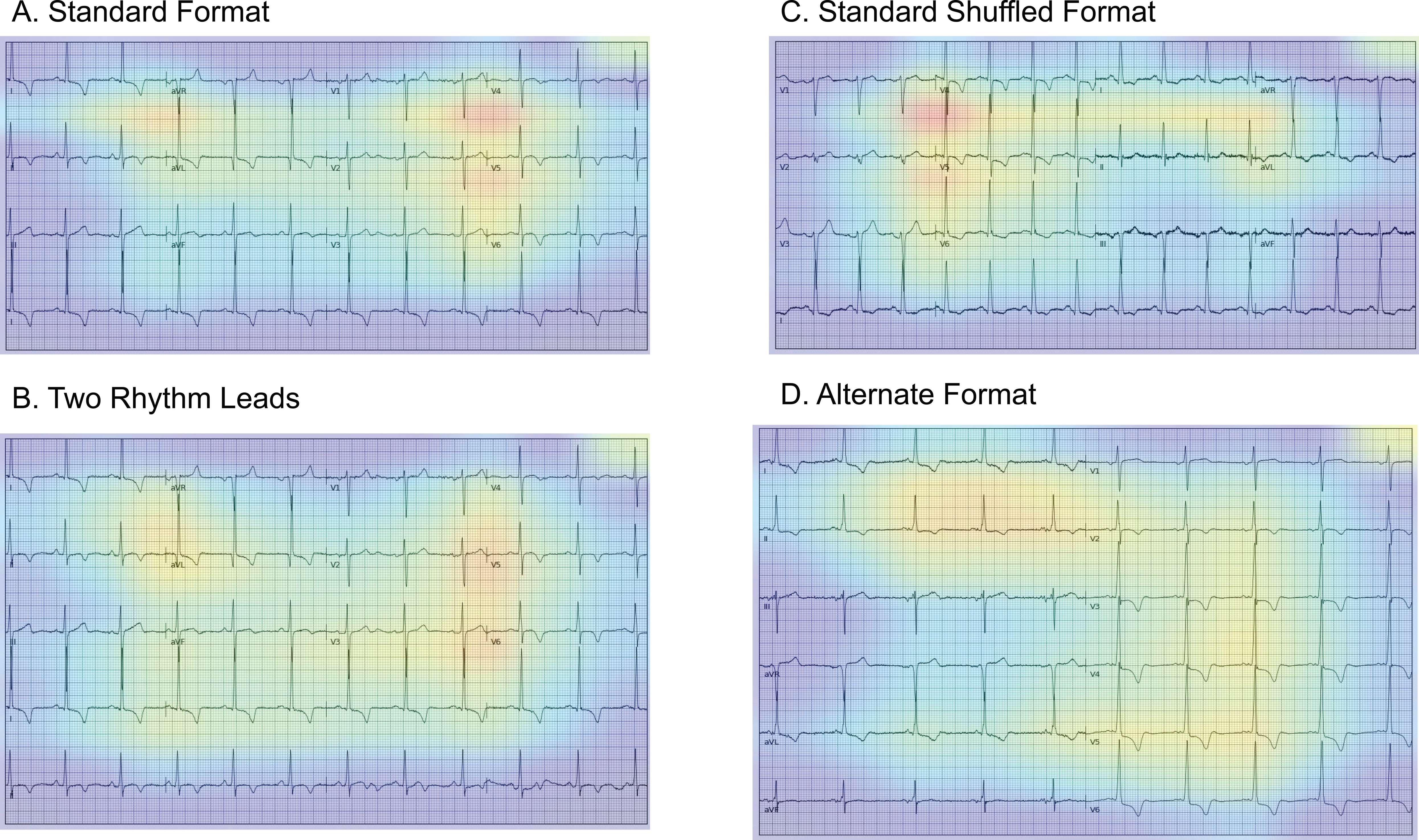
Gradient-weighted Class Activation Mapping (Grad-CAMs) across Electrocardiogram formats. A) Standard format B) Two rhythm leads C) Standard shuffled format D) Alternate format. The heatmaps represent averages of the 25 positive cases with the most confident model predictions for HCM.

We performed a sensitivity analysis to evaluate the model in images plotted using the custom real-world plotting strategy. Specifically, we compared the model to one trained using only the four image formats in **Extended Figure 2** to assess the incremental value of the data augmentation strategy. Of note, while the simple CNN model trained without the custom data augmentation strategy performed well when evaluated on held-out test set images plotted in only the standard format (AUROC 0.959, AUPRC 0.754, F1 0.613), it had a substantial drop-off in performance when tested on images plotted in varied formats using the custom plotting strategy (AUROC 0.924, AUPRC 0.545, F1 0.378). In contrast, our final model demonstrated consistent performance in the test set containing ECG images in varied formats (**Table S2**).

### Localization of Predictive Cues for HCM

We used Gradient-weighted Class Activation Mapping (Grad-CAM) to obtain a heatmap highlighting the portions of ECG images that were important for predicting HCM.^22^ This provides interpretability for the model’s predictions, and allows clinicians to determine whether model predictions are based on clinical information contained in the ECG data, or based on spurious data features. Class activation heatmaps of the 25 positive cases with the most confident model predictions for HCM prediction across four ECG layouts are presented in **Figure 3**. For all four formats of images, the region corresponding to leads V4 and V5 was the most important areas for the prediction of HCM. Representative images of Grad-CAM analysis in sampled individuals with positive screens in UK Biobank, the external validation site, showed similar patterns (**Extended Figure 4**).

**Figure 4.**
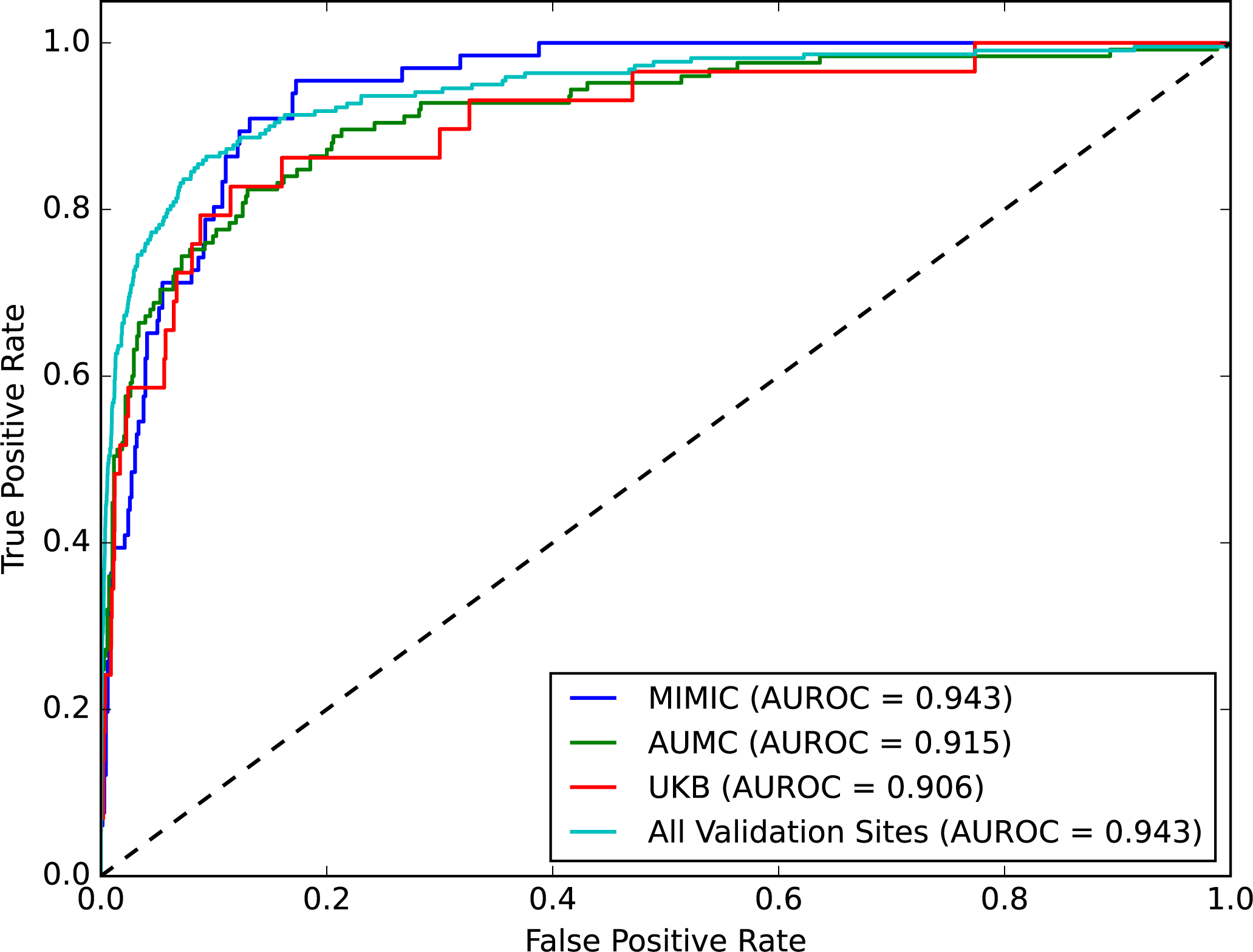
Receiver-Operating Curves for external validation sites. Abbreviations: AUROC, area under receiver-operating characteristic curve; AUMC, Amsterdam University Medical Center; UKB, UK Biobank.

### External Validation

We pursued external validation in a series of clinical and population-based cohort studies. The first of these was in the MIMIC-IV database, comprising of patient-related data collected at the Beth Israel Deaconess Medical Centre (BIDMC) between 2008 and 2019.^23,24^ The dataset is freely accessible to credentialed users. We used inclusion and exclusion criteria in a manner similar to the internal YNHH dataset. Specifically, clinicians adjudicated whether cardiac MRIs indicated HCM, and the nearest ECG within one year of an indicative MRI was included. Controls were age- and sex-matched to HCM ECGs at a 10:1 prevalence. The dataset included 726 ECGs from unique patients, including 66 (9.1%) patients with HCM. The median age of the patients at time of the ECG was 60 years (IQR 52 – 65), and 242 (33.3%) were women (**Extended Table 3**). The model demonstrated an AUROC of 0.94 (0.92-0.96) and an AUPRC of 0.63 (0.51-0.74) in this set.

The second validation dataset derived from consecutive patients undergoing cardiac MRIs at the Amsterdam University Medical Center (AUMC). All cardiac MRIs collected between 2015 and 2022 were reviewed by a cardiologist, and cases were defined as those with a diagnosis of HCM based on the MRI, while controls were defined as patients with normal MRI findings. The nearest ECG from 1 year prior to any time after the cardiac MRI was selected. The AUMC CMR cohort consisted of ECGs from 807 individual patients, 125 (15.5%) of whom were identified to have HCM. Individuals had a median age of 57 years (IQR 46 – 66) at the time of ECG recording, and 386 (47.8%) were women (**Extended Table 3**). The model had an AUROC of 0.92 (0.88 – 0.95) and an AUPRC of 0.77 (0.69 – 0.83).

Finally, we applied the model to the UK Biobank validation set, consisting of 45,389 ECGs from prospectively enrolled individuals, including 29 (0.06%) with an ICD code for HCM. The median age of the patients was 64 years (IQR 58 – 70), and 23,376 (51.5%) were women (**Extended Table 3**). The model had an AUROC of 0.91 (0.89 – 0.99) on these images, with a sensitivity of 0.72 and specificity of 0.92 at the threshold set in the development population. The validation performance of the model was consistent across the 3 validation datasets (**Figure 4, Extended Table 4**). In a mixed sample of data from all external validation sites, the model demonstrated an AUROC and AUPRC of 0.94 (0.92 – 0.96) and 0.33 (0.27 – 0.39), respectively, in detecting HCM.

### Evaluation of LVH Phenotype in False Positives

To assess the appropriateness of our approach as a screening strategy – a high false positive rate is a major concern for low-prevalence, highly underdiagnosed conditions like HCM – we evaluated phenotypic characteristics of false positives. We deployed the model on 5,000 randomly selected ECGs recorded within 30 days of a TTE in patients outside our development and held-out test sets, thereby excluding any patient with known HCM. Of these, 765 (15.3%) were classified as false positives and 4,235 (84.7%) as true negatives. We extracted end-diastolic interventricular septal wall thickness (IVSd) and compared IVSd measurements in false positives and true negatives. The median IVSd among the false-positive subset was 10.8 mm (IQR 9.5 – 12.0), compared with 9.5mm (IQR 8.3 – 10.8) among false negatives (p < 0.001 by Wilcoxon rank sum test) **(Figure 5A)**.

**Figure 5.**
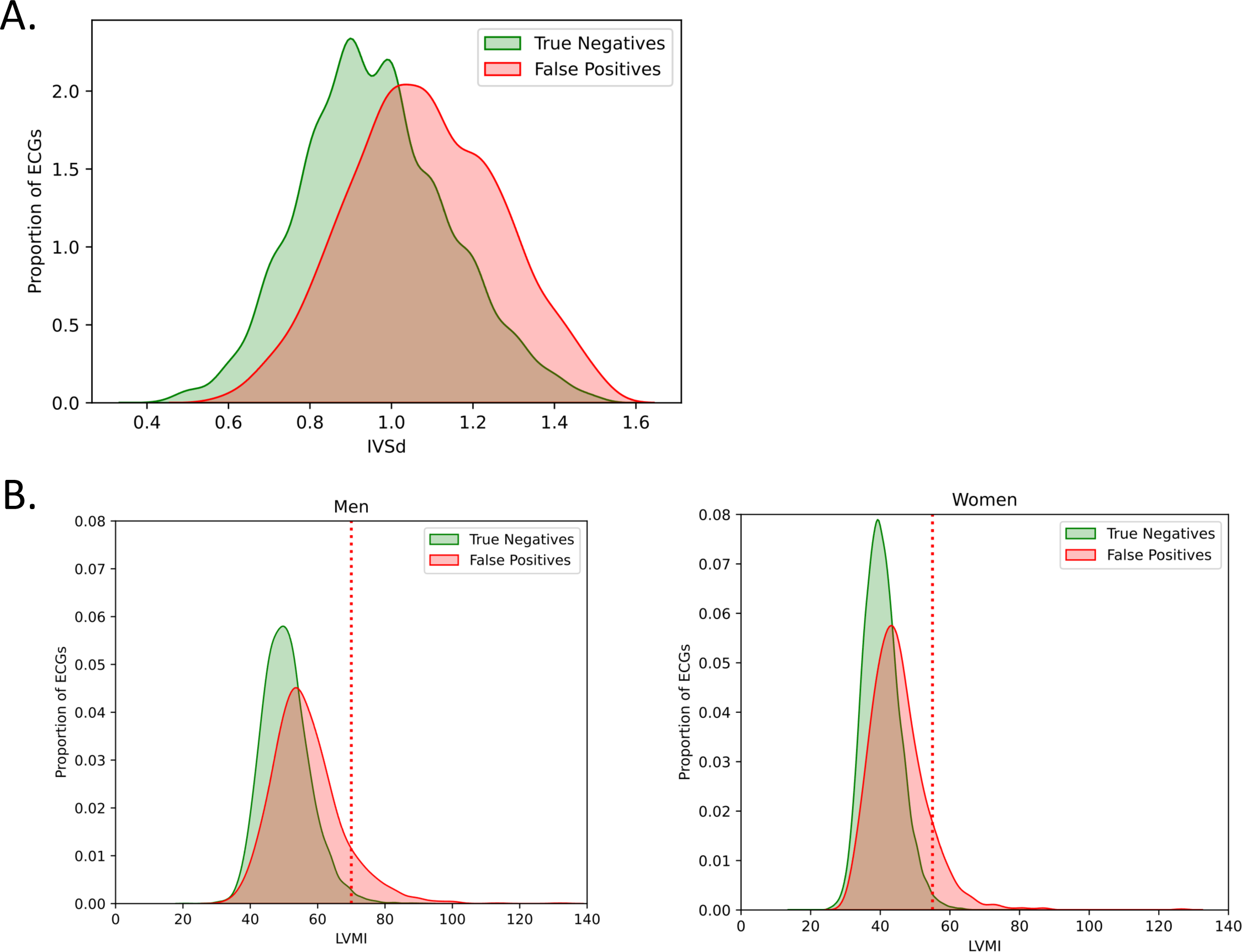
(A) Distribution of interventricular septal thickness in Yale New Haven Hospital patients without hypertrophic cardiomyopathy. (B). Distribution of left ventricular mass index in UK Biobank without hypertrophic cardiomyopathy.

We also performed a similar analysis in UK Biobank. Of the 45,360, ECGs in the UK Biobank validation set without an HCM diagnosis, 36,332 were from individuals who had undergone cardiac MRIs and had recorded measures that could be used to calculate LVMI. The model classified 2,732 (10.3%) as false positives and 33,600 (92.5%) as true negatives. We used cardiac MRI derived left-ventricular mass index (LVMI) to compare the characteristics of participants with positive and negative AI-ECG screens, defining LVH as LVMI > 70 g/m^2^ in men and LVMI > 55 g/m^2^ in women.^25,26^ Of the false positive screens, 282 (7.9%) had LVH, compared with 361 (1.1%) among true negative screens **(Figure 5B)**.

## DISCUSSION

We developed and validated an automated deep-learning model for identifying HCM from ECG images. The model has consistent performance across variations in ECG waveform layouts, making it suitable for implementation in various settings. Moreover, the model has excellent discrimination and sensitivity, which are ideal for screening. The model was developed and tested in a diverse population with high performance in subgroups of age, sex, and race. Moreover, the algorithm was validated and demonstrated consistent performance at three geographically dispersed settings that spanned clinical settings. These settings included MIMIC-IV, which contained ECGs from patients seen at a large academic center in the United States, AUMC, a cardiomyopathy referral center in the Netherlands, and UK Biobank, a population-based cohort, where protocolized electrocardiograms and cardiac MRIs were performed. Class-discriminating signals were localized to the anterior and lateral leads regardless of the ECG layout, topologically corresponding to the left ventricle.

With the emergence of novel therapies for HCM which target the underlying basis for disease expression there is a greater importance for early and reliable diagnosis of HCM. The ability to leverage and ultimately integrate ECG-image-based deep learning models into health care systems represents a novel application of AI that can improve clinical care and public health by offering an easily accessible and highly accurate modality for the disease detection.^18,19,27,28^. Studies show that the traditional interpretation of visible ECG features for HCM has limited accuracy^29,30^ and lacks specificity.^10,31^ Deep learning models utilizing raw ECG voltage signals have been proposed to detect HCM.^8,9,12^ While these models have excellent performance in internal validation, few have been developed using robust clinical criteria for defining HCM, and only a few have been externally validated. In a literature synthesis, our model matches or surpasses the performance of these signal models during both internal and external validation (**Table S3**). It is critical in validating new tools for HCM to incorporate objective advanced imaging-based biologic features to identify disease to ensure the generalizability of deep learning models in diverse settings.^8,18,32^ Our use of CMR to identify disease represents a robust approach for case identification.^8,10^ This validation is also demonstrated by consistently high discrimination of the model for HCM in the patient subgroup with increased septal wall thickness.

The major advantage of this method is that using images to detect HCM allows for the accessible implementation of a potential ECG-based screening approach. Digital or printed ECG images are the most commonly available format. Clinicians can use the model on screenshots or export images of a ECGs in the EHR or capture smartphone photographs of a printed ECGs. While certain health systems subscribe to systems that provide access to ECG signal waveform data, these systems are not universally available, especially in low-resource settings.^33^ Moreover, ECG images are an interoperable data stream not tied to proprietary formats from specific ECG machine vendors, making them readily available to clinicians at the point of care. Further, previous reports, including our recent work, have demonstrated similar performance of AI-ECG models using ECG signals and images.^19,34^

Currently, the guidelines for universal screening for HCM are equivocal, given the limited affordability of advanced cardiac imaging and the high number of false-positive and negative screens on clinically apparent ECG anomalies.^35–37^ However, an accurate and accessible approach to HCM diagnosis using AI can potentially make HCM screening economically viable. Thus, there is an urgent need for prospective clinical and cost-effectiveness assessments for AI-ECG-based HCM screening, especially for people at elevated risk of sudden cardiac death, such as young athletes.^11,38–40^

In addition, using ECG images in our model overcomes the implementation challenges of black box algorithms. The consistent localization of the risk-discriminative signals in anterior and lateral leads of ECG images, regardless of the lead location on printed images, indicates the left ventricular origin of the underlying pathology. Visual representations consistent with clinical knowledge could explain parts of the model prediction process and address the hesitancy in using these tools in clinical practice.^41^

Our study has several limitations. First, our model was developed using retrospective data from a single center, including ECGs from patients with clinical indications for both ECGs and advanced cardiac imaging such as CMR or echocardiography. While the Yale New Haven Health System serves a large and diverse population in Connecticut and Rhode Island, and the model was externally validated in three settings, including the community-based cohort of UK Biobank prospective validation of the models is essential before widespread deployment in a screening setting. Second, despite the model’s high discrimination for HCM, its implementation for community-based screening can result in high false positive rates. Since screening for low prevalence conditions inherently limits the yield for diagnostic modalities, a role for universal HCM screening must be established before widespread deployment of the model.^42,43^ Third, despite localizing the class-discriminative signals in the ECG image to the left ventricular areas, heatmap analysis may not capture all the model predictive features, such as the duration of ECG segments, intervals, or ECG waveform morphologies that might have been used in model predictions. Fourth, the model demonstrated a lower sensitivity and higher specificity on the UK-Biobank cohort, composed of younger and generally healthier individuals. However, we relied on diagnosis codes to identify HCM cases during testing in the UK Biobank. Our analysis of participants screened as false positives showed significantly higher LVMI than true negatives, suggesting that the diagnosis-code-based classification could have missed some cases of HCM. Regardless, depending on the intended result of the screening approach and resource constraints with downstream testing, prediction thresholds for HCM may need to be recalibrated when deployed in such settings. Fifth, 2D ECG images often contain less than 10 seconds of data for all 12 leads. While ECG waveform signal models are trained and deployed on all 10 seconds of data across 12 leads, we found that our model – which is trained and deployed on images that may contain only 2.5 seconds of some leads – has performance that matches or exceeds previously published ECG waveform models. Further, previous reports have demonstrated similar performance for AI-ECG models developed using 10-second ECG signal data for all leads and ECG images plotted in the formats commonly used in clinical care. Sixth, the model demonstrated lower specificity in subsets of ECGs with atrial fibrillation and left bundle branch block. While it is possible that some of the explanatory power of the model is coming from these findings, they are not specific to HCM and the subsets are small in number (n=75 and n=19, respectively). Further, the model discrimination in these subsets is similar to the overall population, with overlapping AUROC confidence intervals.

In conclusion, we developed and validated a high-performing deep learning-based model that detects HCM from images of clinical 12-lead ECGs. This approach represents an accessible strategy for earlier HCM detection, especially in low-resource settings, with the potential to enhance care for patients.

## METHODS

The Yale Institutional Review Board approved the study protocol and waived the need for informed consent as the study represents a secondary analysis of existing data. Patients who opted out of research studies at the Yale New Haven Hospital (YNHH) were excluded. An online version of the model is publicly available for research use at https://www.cards-lab.org/ecgvision-hcm. This web application represents a prototype of the eventual application of the model, with instructions for required image standards and a version that demonstrates an automated image standardization pipeline.

### Data Source and Study Population

We used 12-lead ECG signal waveform data collected during the clinical care of patients at the YNHH between 2012 and 2021. These ECGs were recorded as standard 12-lead recordings sampled at 500 Hz for 10 seconds. They were recorded on multiple different machines, with Philips PageWriter and GE MAC the most frequently used.

We identified the earliest MRI reports between 2013 and 2022 for 1,061 patients containing any mention of HCM. Each report was manually reviewed by at least two of three reviewing clinicians (LSD, AA, EKO), with any disagreements resolved by a fourth clinical reviewer (RK). In total, 904 people were identified as having confirmed or possible HCM. Of these, 779 reports included a confirmed HCM diagnosis and 125 were identified as possible HCM, defined as the inclusion of HCM as one of the reported differential diagnoses or the presence of features potentially suggestive, but not conclusive for HCM. The data from these patients were split into mutually exclusive training, validation, and test sets in an 85:5:10 ratio. Only MRI-confirmed HCM cases were retained in validation and testing, while confirmed and possible HCM cases were included in the training cohort. This ensured the model was exposed to a broad spectrum of HCM phenotypes during training, while maintaining the specificity and robustness of the diagnostic labels during validation and testing.

For each of these patients, all ECGs recorded up to a year before the MRI and any time after the MRI was considered, except for those ECGs after a septal reduction procedure, including alcohol septal ablation or ventricular myectomy. To further augment the training cohort, we incorporated ECGs from patients whose transthoracic echocardiograms (TTE) demonstrated severe ventricular hypertrophy (LVH), defined by interventricular septal thickness in diastole (IVSd) of greater than 15 mm. We posited that these may represent individuals with possible HCM. ECGs performed within 30 days before or after a TTE demonstrating severe LVH were included as cases in the training set but were not considered cases in the test set, which only included CMR-confirmed HCM. This further broadened the comprehensiveness of the HCM phenotype during model development.

For patients with more than one recorded ECG filling these criteria, a maximum of five most recent ECGs were used to create cohorts to avoid overrepresenting patients undergoing frequent ECGs. To derive a control cohort for the training set, we identified ECGs recorded within 30 days of a TTE in a cohort of patients who did not have a diagnosis code recorded for HCM (**Table S4**) and were not in the cohort of patients with possible positive HCM cases based on MRI reports or IVSd values. Thus, the control cohort did not include patients with any ICD code for HCM (**Table S4**), any mention of HCM in cardiac MRI reports, or any TTE with IVSd > 15mm. We randomly sampled control ECGs, so the train set had a 1:9 ratio of HCM ECGs to controls, which allowed the model to learn the signatures of HCM on ECGs successfully.

For the validation and test sets, controls represented ECGs from individuals age-/sex-matched to HCM cases (10:1), excluding those who had a hospitalization with heart failure with a diagnosis code suggestive of cardiomyopathy **(Table S4)**. Age- and sex-matching is essential as HCM more commonly affects men and is diagnosed in the younger population, requiring the confirmation that age and sex differences in the HCM population do not drive the presumed HCM signature learned from ECGs. To implement this, we randomly selected 10 control patients within 5 years of age at the time of the selected ECG and of the same sex as each HCM case. We ensured no patient overlap between training, validation, and test sets.

### Image Generation

We generated ECG images to recapitulate the variation in ECG layouts in a real-world setting. Our approach to image plotting has been previously described and represents the processing steps of ECG machines to convert acquired waveform data to printed outputs.^15–19^ All ECGs were analyzed to determine whether they had 10 seconds of continuous recordings across all 12 leads. The 10-second samples were preprocessed with a one-second median filter, subtracted from the original waveform to remove baseline drift in each lead. Converting ECG signals to images was independent of model development, ensuring that the model did not learn any aspects of the processing that generated images from the signals.

ECG signals were transformed into ECG images using custom plotting software. Images were generated with a calibration of 10 mm/mV, which is standard for printed ECGs in most real-world settings. Using the Python Image Library (PIL v9.2.0), we converted all images to greyscale, then down-sampling to 300×300 pixels regardless of their original resolution. Given that real-world ECG images may vary in the layout of leads, we created a dataset with several different plotting schemes for each signal waveform recording (**Figure 1**). Variations included but were not limited to the format of the plotted ECG, colors the original ECG was plotted in before conversion to grayscale, lead label font, size and position, and grid and signal line width. Plotted formats included the four formats used in previous studies – standard (four columns printed sequentially with each containing 2.5-second intervals from three leads as well as a rhythm strip), two-rhythm (a second rhythm lead added), alternate (two columns with simultaneous 5-second recordings in each), and shuffled (precordial leads in the first two columns and limb leads in the third and fourth).^18^ Additionally, formats with zero and three rhythm leads were plotted. Leads I, II, V1, and V5 were randomly selected as rhythm leads for images where rhythm leads were present. Examples of figures used during training are included in **Extended Figure 1**. For evaluation, ECG images were plotted in the four formats used in previous studies (**Extended Figure 2**). All images were rotated a random amount between -10 and 10 degrees before input into the model. In a sensitivity analysis, we evaluated the model in images plotted using the custom plotting strategy, encompassing variations seen in real-world settings. This model was compared with a simpler model trained using the four image formats to assess the incremental value of the data augmentation strategy.

### Model Architecture and Training

We built a convolutional neural network model based on the EfficientNet-B3 architecture.^20^ The EfficientNet-B3 model requires images to be sampled at 300 x 300 square pixels, includes 384 layers, and has over 10 million trainable parameters. To allow label-efficient model development, we initialized the model using weights from a pretrained EfficientNet-B3 model that leveraged a novel self-supervised biometric contrastive learning approach, wherein the model was trained to identify individual patient-specific patterns in ECGs regardless of their interpretation.^21^ None of the ECGs on the self-supervised pretraining task represented individuals in the model development. For training, we first unfroze the last four layers and trained the model with a learning rate of 0.01 for 2 epochs. Then, we unfroze all layers and trained the model with a learning rate of 5 × 10^−6^ for 6 epochs. We used an Adam optimizer, gradient clipping, and a minibatch size of 64 throughout training. The optimizer and learning rates were chosen to balance training time and performance. For both stages of training the model, we stopped training when validation loss did not improve in 3 consecutive epochs. A custom class-balanced loss function (weighted binary cross-entropy) based on the effective number of samples was used given that the case and control labels were not equally balanced.

### Localization of Model Predictive Cues

To obtain a heatmap highlighting the portions of an ECG image that were important for predicting HCM, we used Gradient-weighted Class Activation Mapping (Grad-CAM).^22^ We calculated the gradients on the final stack of filters in our EfficientNet-B3 model for each prediction. We performed a global average pooling of the gradients in each filter, emphasizing those that contributed to a prediction. We then multiplied these filters by their importance weights and combined them across filters to generate Grad-CAM heatmaps. Among the 25 positive cases with the most confident model predictions for HCM across ECG formats, we averaged class activation maps to determine the most important image areas for the prediction of HCM. We took an arithmetic mean across the heatmaps for a given image format and overlayed this average heatmap across a representative ECG before converting the image to grayscale. The Grad-CAM intensities were converted from their original scale (0 – 1) to a color range using the jet colormap array in the Python library matplotlib, which was then overlaid on the original ECG image with an alpha of 0.3. The activation map, a 10×10 array, was upsampled to the original image size using the bilinear interpolation built into TensorFlow v2.8.0. We also evaluated the Grad-CAM for individual ECGs in the UK Biobank to evaluate the consistency of the information on individual examples.

### External Validation

For external, validation we performed a series of clinical and population-based cohort studies. These included (1) the MIMIC-IV database, (2) consecutive patients undergoing cardiac MRIs at the Amsterdam University Medical Center (AUMC) and (3) UK Biobank, a prospective cohort.

MIMIC-IV comprises of patient-related data collected at the Beth Israel Deaconess Medical Centre (BIDMC) between 2008 and 2019.^23,24^ This public-access database is freely accessible to any credentialed PhysioNet user.^44^ Inclusion and exclusion criteria for this set were similar to the internal YNHH set, with clinician-adjudication of cardiac MRIs. Patients were limited to those having a 12-lead ECG within one year of the MRI, and the nearest ECG to the MRI was considered. For controls, we considered ECGs from all patients other than the cohort that had confirmed or possible HCM cases based on MRI reports. The controls were age- and sex-matched to HCM ECGs at a 10:1 prevalence.

Next, we used 12-lead signal waveform data collected during clinical care of patients at the Amsterdam UMC (AUMC) who underwent cardiac MRIs between 2015 and 2022. All cardiac MRIs were reviewed by a cardiologist, with their determination serving as the label for confirmed HCM. Controls were defined as patients with normal cardiac MRI findings. All ECGs recorded between 2012 and 2022 were extracted, and the nearest ECG from 1 year prior to any time after the cardiac MRI was selected. ECGs were recorded as standard 12-lead 10-second recordings collected from Philips and GE machines and were resampled to 500 Hz before plotting.

Finally, we used data from the UK Biobank, under research application #71033, to pursue external validation of our model. UK Biobank represents the largest population-based cohort of 502,468 people in the United Kingdom with protocolized imaging and laboratory testing, along with linked electronic health records. We used linked electronic health records for the participants to identify the presence of HCM diagnosis codes and evaluated our model on all available ECG recordings. In patients without HCM, we also used CMR-derived left-ventricular mass index (LVMI) to compare the characteristics of participants with a positive and negative AI-ECG screen for HCM. LVH was defined as LVMI > 70 g/m^2^ in men and LVMI > 55 g/m^2^ in women.^25,26^

### Evaluation of Hypertrophy in Patients without Confirmed HCM

A high false positive rate or type 1 error is a major concern for screening strategies for low-prevalence conditions like HCM. Furthermore, given that HCM is often underdiagnosed, evaluating the phenotypic characteristics of false positive cases is important. Therefore, among patients without CMR-confirmed HCM, we applied the model to 5,000 randomly selected ECGs recorded within 30 days of a TTE. These ECGs were taken from patients previously not analyzed in development or evaluation sets. We extracted the end-diastolic interventricular septal wall thickness (IVSd), which is available as a continuous measure from TTEs. To evaluate the instances of model-positive screens in patients without CMR-confirmed HCM, we compared the IVSd measurements in false positive and true negative screens.

### Statistical Analysis

Categorical variables were reported as number (percentage, %), and continuous variables as mean (standard deviation [SD]) or median (interquartile range [IQR]), as appropriate. The model’s performance was presented as area under the receiver operating characteristic curve (AUROC) and area under the precision-recall curve (AUPRC). The 95% confidence intervals (CI) for AUROC and AUPRC were calculated using DeLong’s algorithm and bootstrapping with 1000 iterations, respectively.^45,46^ Furthermore, we reported sensitivity, specificity, positive predictive value (PPV), negative predictive value (NPV), and F1 score of the model at the model threshold for 90% sensitivity in the validation set. We also evaluated the longitudinal pattern of the AI-ECG HCM phenotype in all ECGs from patients with HCM in the held-out test set.

Further, the IVSd values in false positive and true negative patients were compared using the Wilcoxon rank sum test. All statistical tests were 2-sided, and the significance level was 0.05. Analytic packages used in model development and statistical analysis are reported in **Table S5**.

## Data Availability

The dataset is not publicly available because they are electronic health records. Sharing this data externally without proper consent could compromise patient privacy and violate the Institutional Review Board’s approval for the study. However, de-identified data may be made available upon reasonable request to the corresponding author, subject to institutional and ethical approvals by the Review Board.

## Code Availability

The code for the study is available at https://github.com/CarDS-Yale/AI_ECG_HCM_Code_Share.

## Author Contributions Statement

V.S., L.S.D, and R.K. conceived and designed the study. V.S. and L.S.D. conducted statistical analyses. V.S, L.S.D., A.A., P.M.C., E.K.O., and R.K. interpreted the data. V.S. and L.S.D. wrote the first draft of the paper and prepared figures. A.A., P.M.C., N.V.S., S.S., M.W.M., M.S.M., H.M.K., F.W.A., E.K.O., and R.K. critically revised the paper. R.K. acquired funding and supervised the study. All authors had full access to all data used in this study. All authors approved the final version for submission.

## REFERENCES

1. Maron, B. J. Hypertrophic cardiomyopathy. JAMA 287, 1308–1320 (2002).

2. Maron, B. J. & Maron, M. S. Hypertrophic cardiomyopathy. Lancet 381, 242–255 (2013).

3. Semsarian, C., Ingles, J., Maron, M. S. & Maron, B. J. New perspectives on the prevalence of hypertrophic cardiomyopathy. J. Am. Coll. Cardiol. 65, 1249–1254 (2015).

4. Sabater-Molina, M., Pérez-Sánchez, I., Hernández del Rincón, J. P. & Gimeno, J. R. Genetics of hypertrophic cardiomyopathy: A review of current state. Clin. Genet. 93, 3–14 (2018).

5. Geske, J. B., Ommen, S. R. & Gersh, B. J. Hypertrophic cardiomyopathy. JACC Heart Fail. 6, 364–375 (2018).

6. Maron, B. J. et al. Diagnosis and evaluation of hypertrophic cardiomyopathy. J. Am. Coll. Cardiol. 79, 372–389 (2022).

7. Writing Committee Members et al. 2020 AHA/ACC guideline for the diagnosis and treatment of patients with hypertrophic cardiomyopathy. Circulation 142, (2020).

8. Goto, S. et al. Multinational Federated Learning Approach to Train ECG and Echocardiogram Models for Hypertrophic Cardiomyopathy Detection. Circulation 146, 755–769 (2022).

9. Ko, W.-Y. et al. Detection of Hypertrophic Cardiomyopathy Using a Convolutional Neural Network-Enabled Electrocardiogram. J. Am. Coll. Cardiol. 75, 722–733 (2020).

10. Finocchiaro, G. et al. The electrocardiogram in the diagnosis and management of patients with hypertrophic cardiomyopathy. Heart Rhythm 17, 142–151 (2020).

11. McLeod, C. J. et al. Outcome of patients with hypertrophic cardiomyopathy and a normal electrocardiogram. J. Am. Coll. Cardiol. 54, 229–233 (2009).

12. Siontis, K. C. et al. Detection of hypertrophic cardiomyopathy by an artificial intelligence electrocardiogram in children and adolescents. Int. J. Cardiol. 340, 42–47 (2021).

13. Magnusson, P., Palm, A., Branden, E. & Mörner, S. Misclassification of hypertrophic cardiomyopathy: validation of diagnostic codes. Clin. Epidemiol. 9, 403–410 (2017).

14. Farahani, N. Z. et al. Explanatory analysis of a machine learning model to identify hypertrophic cardiomyopathy patients from EHR using diagnostic codes. Proceedings (IEEE Int. Conf. Bioinformatics Biomed.) 2020, 1932–1937 (2020).

15. Daskalov, I. K., Dotsinsky, I. A. & Christov, I. I. Developments in ECG acquisition, preprocessing, parameter measurement, and recording. IEEE Eng. Med. Biol. Mag. 17, 50–58 (1998).

16. Blanco-Velasco, M., Weng, B. & Barner, K. E. ECG signal denoising and baseline wander correction based on the empirical mode decomposition. Comput. Biol. Med. 38, 1–13 (2008).

17. Sangha, V. et al. Automated multilabel diagnosis on electrocardiographic images and signals. Nat Commun 13, 1583 (2022).

18. Sangha, V. et al. Detection of Left Ventricular Systolic Dysfunction From Electrocardiographic Images. Circulation (2023) doi:10.1161/CIRCULATIONAHA.122.062646.

19. Dhingra, L. S. et al. Ensemble deep learning algorithm for structural heart disease screening using electrocardiographic images: PRESENT SHD. J. Am. Coll. Cardiol. (2025) doi:10.1016/j.jacc.2025.01.030.

20. Mingxing Tan and Quoc V Le. EfficientNet: Rethinking model scaling for convolutional neural networks. International Conference on Machine Learning, 2019.

21. Sangha, V. et al. Biometric contrastive learning for data-efficient deep learning from electrocardiographic images. J. Am. Med. Inform. Assoc. (2024) doi:10.1093/jamia/ocae002.

22. Selvaraju, R. R. et al. Grad-CAM: Visual Explanations from Deep Networks via Gradient-based Localization. 2017 Ieee International Conference on Computer Vision (Iccv) 618–626 (2017).

23. Gow, B. et al. MIMIC-IV-ECG: Diagnostic electrocardiogram matched subset. PhysioNet 10.13026/4NQG-SB35 (2023).

24. Johnson, A., Pollard, T., Horng, S., Celi, L. A. & Mark, R. MIMIC-IV-Note: Deidentified free-text clinical notes. PhysioNet 10.13026/1N74-NE17 (2023).

25. Petersen, S. E. et al. Reference ranges for cardiac structure and function using cardiovascular magnetic resonance (CMR) in Caucasians from the UK Biobank population cohort. J. Cardiovasc. Magn. Reson. 19, (2017).

26. Naderi, H. et al. Predicting left ventricular hypertrophy from the 12-lead electrocardiogram in the UK Biobank imaging study using machine learning. Eur. Heart J. Digit. Health 4, 316–324 (2023).

27. Oikonomou, E. K. et al. Artificial intelligence-enhanced risk stratification of cancer therapeutics-related cardiac dysfunction using electrocardiographic images. Circ. Cardiovasc. Qual. Outcomes (2024) doi:10.1161/CIRCOUTCOMES.124.011504.

28. Dhingra, L. S. et al. Heart failure risk stratification using artificial intelligence applied to electrocardiogram images: a multinational study. Eur. Heart J. (2025) doi:10.1093/eurheartj/ehae914.

29. Delcrè, S. D. L. et al. Relationship of ECG findings to phenotypic expression in patients with hypertrophic cardiomyopathy: A cardiac magnetic resonance study. Int. J. Cardiol. 167, 1038–1045 (2013).

30. Charron, P. et al. Diagnostic value of electrocardiography and echocardiography for familial hypertrophic cardiomyopathy in a genotyped adult population. Circulation 96, 214–219 (1997).

31. Sharma, S. et al. International recommendations for electrocardiographic interpretation in athletes. Eur. Heart J. 39, 1466–1480 (2018).

32. Dhingra, L. S. et al. A multicenter evaluation of the impact of therapies on deep learning-based electrocardiographic hypertrophic cardiomyopathy markers. Am. J. Cardiol. 237, 35–40 (2024).

33. Lamichhane, B. & Neupane, N. Improved healthcare access in low-resource regions: A review of technological solutions. arXiv [cs.CY] (2022).

34. Sau, A. et al. A comparison of artificial intelligence-enhanced electrocardiography approaches for prediction of time-to-mortality using electrocardiogram images. Eur. Heart J. Digit. Health (2024) doi:10.1093/ehjdh/ztae090.

35. Norrish, G. et al. Yield of clinical screening for hypertrophic cardiomyopathy in child first-degree relatives. Circulation 140, 184–192 (2019).

36. Lafreniere-Roula, M. et al. Family screening for hypertrophic cardiomyopathy: Is it time to change practice guidelines? Eur. Heart J. 40, 3672–3681 (2019).

37. Corrado, D., Basso, C., Schiavon, M. & Thiene, G. Screening for hypertrophic cardiomyopathy in young athletes. N. Engl. J. Med. 339, 364–369 (1998).

38. Screening program in the athlete in the suspicion of HCM. J. Integr. Cardiol. 1, (2015).

39. Anderson, B. R., McElligott, S., Polsky, D. & Vetter, V. L. Electrocardiographic screening for hypertrophic cardiomyopathy and long QT syndrome: The drivers of cost-effectiveness for the prevention of sudden cardiac death. Pediatr. Cardiol. 35, 323–331 (2014).

40. Aminorroaya, A. et al. Development and multinational validation of an ensemble deep learning algorithm for detecting and predicting structural heart disease using noisy single-lead electrocardiograms. Eur. Heart J. Digit. Health (2025) doi:10.1093/ehjdh/ztaf034.

41. Albahri, A. S. et al. A systematic review of trustworthy and explainable artificial intelligence in healthcare: Assessment of quality, bias risk, and data fusion. Inf. Fusion 96, 156–191 (2023).

42. O’Toole, B. I. Screening for low prevalence disorders. Aust. N. Z. J. Psychiatry 34, A39–A46 (2000).

43. Basavarajaiah, S. et al. Prevalence of hypertrophic cardiomyopathy in highly trained athletes. J. Am. Coll. Cardiol. 51, 1033–1039 (2008).

44. Goldberger, A. L. et al. PhysioBank, PhysioToolkit, and PhysioNet: components of a new research resource for complex physiologic signals. Circulation 101, E215–20 (2000).

45. Sun, X. & Xu, W. Fast implementation of DeLong’s algorithm for comparing the areas under correlated receiver operating characteristic curves. IEEE Signal Process. Lett. 21, 1389–1393 (2014).

46. DeLong, E. R., DeLong, D. M. & Clarke-Pearson, D. L. Comparing the areas under two or more correlated receiver operating characteristic curves: a nonparametric approach. Biometrics 44, 837–845 (1988).

