## Supplementary material for "Identification of Hypertrophic Cardiomyopathy on Electrocardiographic Images with Deep Learning": Online Supplement

**Table of Contents**

**Table S1 | Performance of model on test images across age ranges in the age sex matched held-out test set ……………………..….2**

**Table S2 | Comparison of models trained with and without data augmentation on electrocardiograms in the held-out test set plotted in only standard image format and plotted with data augmentations ……………………………………………………3**

**Table S3 | Comparison of model performance on internal held-out test set and external validation datasets with reported performances of two previously published signal-based 1D Convolutional Neural Network models ……………………………..4**

**Table S4 | International classification of disease tenth revision (ICD-10) codes for hypertrophic cardiomyopathy and conditions indicating cardiomyopathies ………………………………………………………………………………………5**

**Table S1. Performance of model on test images across age ranges in the age sex matched held-out test set.** Abbreviations: TP, True Positives; TN, True Negatives; FP, False Positives; FN, False Negatives; AUROC, area under receiver operating characteristic curve; AUPRC, area under precision recall curve.

| Age Range | TP | TN | FP | FN | Specificity | Sensitivity | AUROC | AUPRC | F1 Score |
|---|---|---|---|---|---|---|---|---|---|
| **All** | 133 | 1349 | 151 | 17 | 0.899 | 0.887 | 0.949 | 0.741 | 0.613 |
| **18-30** | 4 | 40 | 0 | 0 | 1 | 1 | 1 | 1 | 1 |
| **31-40** | 19 | 234 | 16 | 6 | 0.936 | 0.76 | 0.918 | 0.743 | 0.633 |
| **41-50** | 15 | 151 | 9 | 1 | 0.944 | 0.938 | 0.989 | 0.918 | 0.75 |
| **51-60** | 37 | 366 | 34 | 3 | 0.915 | 0.925 | 0.962 | 0.835 | 0.667 |
| **61-70** | 33 | 322 | 48 | 4 | 0.87 | 0.892 | 0.952 | 0.642 | 0.559 |
| **71-80** | 19 | 191 | 29 | 3 | 0.868 | 0.864 | 0.943 | 0.634 | 0.543 |
| **81-90** | 6 | 45 | 15 | 0 | 0.75 | 1 | 0.894 | 0.641 | 0.444 |

**Table S2. Comparison of models trained with and without data augmentation on electrocardiograms in the held-out test set plotted in only standard image format and plotted with data augmentations.**

| Model Type | Test Images | PPV | NPV | Specificity | Sensitivity | AUROC | AUPRC | F1 Score |
|---|---|---|---|---|---|---|---|---|
| Data Augmentation | Standard Images | 0.468 | 0.988 | 0.899 | 0.887 | 0.949 (0.929-0.969) | 0.741 (0.664-0.803) | 0.613 |
| | Varied Images | 0.438 | 0.991 | 0.882 | 0.92 | 0.946 (0.924-0.968) | 0.735 (0.664-0.805) | 0.594 |
| No Data Augmentation | Standard Images | 0.511 | 0.99 | 0.913 | 0.907 | 0.959 (0.941-0.977) | 0.754 (0.681-0.829) | 0.654 |
| | Varied Images | 0.235 | 0.994 | 0.688 | 0.96 | 0.924 (0.904-0.944) | 0.545 (0.473-0.644) | 0.378 |

**Table S3. Comparison of model performance on internal held-out test set and external validation datasets with reported performances of two previously published signal-based 1D Convolutional Neural Network models.**

| | | **Sangha et al** | Ko et al | Goto et al |
|---|---|---|---|---|
| Internal Validation | AUROC | 0.95 | 0.96 | 0.9 |
| | Sensitivity | 0.89 | 0.87 | 0.86 |
| | Specificity | 0.9 | 0.9 | 0.8 |
| External Validation | AUROC | 0.94 | 0.92 | 0.93 |
| | Sensitivity | 0.85 | 0.83 | 0.9 |
| | Specificity | 0.92 | 0.88 | 0.78 |

**Table S4. International classification of disease tenth revision (ICD-10) codes for hypertrophic cardiomyopathy and conditions indicating cardiomyopathies.** Note: Conditions indicating cardiomyopathies were only considered as present, if the patients also had an inpatient hospitalization with a heart failure diagnosis code.

| Condition | International Classification of Disease Tenth Revision (ICD-10) Codes |
|---|---|
| **Hypertrophic Cardiomyopathy** | 'I42.1', 'I42.2' |
| **Heart Failure** | 'I50' , 'I50.1' , 'I50.2', 'I50.20', 'I50.21', 'I50.22', 'I50.23', 'I50.3', 'I50.30', 'I50.31', 'I50.32', 'I50.33', 'I50.4', 'I50.40', 'I50.41', 'I50.42', 'I50.43', 'I50.8', 'I50.81' , 'I50.810', 'I50.811', 'I50.812', 'I50.813', 'I50.814', 'I50.82', 'I50.83', 'I50.84', 'I50.89', 'I50.9' |
| **Conditions Indicating Cardiomyopathies** (considered in patients with inpatient hospitalization with a heart failure diagnosis) | 'I21.01', 'I21.02', 'I21.09', 'I21.11', 'I21.19', 'I21.21', 'I21.29', 'I21.3', 'I21.4', 'I21.9', 'I22.0', 'I22.1', 'I22.2', 'I22.8', 'I22.9', 'I23.0', 'I23.1', 'I23.2', 'I23.3', 'I23.4', 'I23.5', 'I23.6', 'I23.7', 'I23.8', 'I24.0', 'I24.8', 'I24.9', 'I25.10', 'I25.110', 'I25.111', 'I25.118', 'I25.119', 'I25.2', 'I25.5', 'I25.5', 'I25.6', 'I25.700', 'I25.701', 'I25.708', 'I25.709', 'I25.710', 'I25.711', 'I25.718', 'I25.719', 'I25.720', 'I25.721', 'I25.728', 'I25.729', 'I25.730', 'I25.731', 'I25.738', 'I25.739', 'I25.750', 'I25.751', 'I25.758', 'I25.759', 'I25.760', 'I25.761', 'I25.768', 'I25.769', 'I25.790', 'I25.791', 'I25.798', 'I25.799', 'I25.810', 'I25.811', 'I25.812', 'I25.82', 'I25.83', 'I25.84', 'I25.89', 'I25.9', 'T82.855A', 'T82.01XA', 'T82.01XD', 'T82.01XS', 'T82.02XA', 'T82.02XD', 'T82.02XS', 'T82.03XA', 'T82.03XD', 'T82.03XS', 'T82.09XA', 'T82.09XD', 'T82.09XS', 'T82.221A', 'T82.221D', 'T82.221S', 'T82.222A', 'T82.222D', 'T82.222S', 'T82.223A', 'T82.223D', 'T82.223S', 'T82.228A', 'T82.228D', 'T82.228S', 'T82.6XXA', 'T82.6XXD', 'T82.6XXS', 'Z95.2', 'Z95.2', 'Z95.3', 'Z95.3', 'Z95.4', 'Z95.4', 'I05.8', 'I05.9', 'I06.8', 'I06.9', 'I07.8', 'I07.9', 'I08.0', 'I08.1', 'I08.2', 'I08.3', 'I08.8', 'I08.9', 'I09.1', 'I34.0', 'I34.1', 'I34.2', 'I34.8', 'I34.9', 'I35.0', 'I35.1', 'I35.2', 'I35.8', 'I35.9', 'I36.0', 'I36.1', 'I36.2', 'I36.8', 'I36.9', 'I37.0', 'I37.1', 'I37.2', 'I37.8', 'I37.9', 'I38.', 'I39.', 'Q22.0', 'Q22.1', 'Q22.2', 'Q22.3', 'Q22.8', 'Q22.9', 'Q23.0', 'Q23.1', 'Q23.8', 'Q23.9', 'Q25.3', 'Q64.2', 'I47.0', 'I47.1', 'I47.2', 'I47.9', 'I48.0', 'I48.1', 'I48.11', 'I48.19', 'I48.2', 'I48.20', 'I48.21', 'I48.3', 'I48.4', 'I48.91', 'I48.92', 'I49.01', 'I49.02', 'I49.1', 'I49.2', 'I49.3', 'I49.40', 'I49.49', 'I49.5', 'I49.8', 'I49.9', ' Z8.6.79', 'I11.0', 'I11.0', 'I11.9', 'I12.0', 'I12.0', 'I12.9', 'I12.9', 'I13.0', 'I13.0', 'I13.0', 'I13.10', 'I13.10', 'I13.11', 'I13.11', 'I13.2', 'I13.2', 'I13.2', 'I16.0', 'I16.1', 'I16.9', 'I67.4', 'I67.4', 'H35.031', 'H35.031', 'H35.032', 'H35.032', 'H35.033', 'H35.033', 'H35.039', 'I42.6', 'I42.6', 'K29.20', 'K29.21', 'K70.40', 'K70.41', 'K70.0', 'K70.2', 'K70.30', 'K70.31', 'K70.9', 'K85.2', 'K85.20', 'K85.21', 'K85.22', 'K86.0', 'K29.21', 'T40.5X1A', 'T40.5X2A', 'T40.5X3A', 'T40.5X4A', 'T51.8X1A', 'T51.8X2A', 'T51.8X3A', 'T51.8X4A', 'T51.91XA', 'T51.92XA', 'T51.93XA', 'T51.94XA', 'T40.5X1A', 'T51.8X1A', 'T51.91XA', 'T40.5X2A', 'T51.8X2A', 'T51.92XA', 'T40.5X3A', 'T51.8X3A', 'T51.93XA', 'T40.5X4A', 'T51.8X4A', 'T51.94XA', 'Y90.2', 'Y90.3', 'Y90.4', 'Y90.5', 'Y90.6', 'Y90.7', 'Y90.8', 'Y90.9', 'Z71.41', 'Z71.42', 'Z63.72', 'K70.10', 'K70.11', 'T40.5X1A', 'T40.5X2A', 'T40.5X3A', 'T40.5X4A', 'T51.8X1A', 'T51.8X2A', 'T51.8X3A', 'T51.8X4A', 'T51.91XA', 'T51.92XA', 'T51.93XA', 'T51.94XA', 'T40.5X5A', 'T40.5X6A', 'T40.5X1D', 'T40.5X2D', 'T40.5X3D', 'T40.5X4D', 'T51.8X1D', |

| | |
|---|---|
| | 'T51.8X2D', 'T51.8X3D', 'T51.8X4D', 'T51.91XD', 'T51.92XD', 'T51.93XD', 'T51.94XD', 'T40.5X5D', 'T40.5X6D', 'T40.5X1S', 'T40.5X2S', 'T40.5X3S', 'T40.5X4S', 'T40.5X5S', 'T40.5X6S', 'T51.8X1S', 'T51.8X2S', 'T51.8X3S', 'T51.8X4S', 'T51.91XS', 'T51.92XS', 'T51.93XS', 'T51.94XS', 'Q86.0', 'F10.150', 'F10.151', 'F10.159', 'F10.250', 'F10.251', 'F10.259', 'F10.950', 'F10.951', 'F10.959', 'F14.150', 'F14.151', 'F14.159', 'F14.250', 'F14.251', 'F14.259', 'F14.950', 'F14.951', 'F14.959', 'F10.180', 'F10.280', 'F10.980', 'F14.180', 'F14.280', 'F14.980', 'T40.5X2A', 'T51.8X2A', 'T51.92XA', 'F10.10', 'F10.120', 'F10.121', 'F10.129', 'F10.130', 'F10.131', 'F10.132', 'F10.139', 'F10.14', 'F10.150', 'F10.151', 'F10.159', 'F10.180', 'F10.181', 'F10.182', 'F10.188', 'F10.19', 'F10.20', 'F10.220', 'F10.221', 'F10.229', 'F10.230', 'F10.231', 'F10.232', 'F10.239', 'F10.24', 'F10.250', 'F10.251', 'F10.259', 'F10.26', 'F10.27', 'F10.280', 'F10.281', 'F10.282', 'F10.288', 'F10.29', 'F10.920', 'F10.921', 'F10.929', 'F10.930', 'F10.931', 'F10.932', 'F10.939', 'F10.94', 'F10.950', 'F10.951', 'F10.959', 'F10.96', 'F10.97', 'F10.980', 'F10.981', 'F10.982', 'F10.988', 'F10.99', 'G31.2', 'G62.1', 'K29.20', 'K29.21', 'K70.0', 'K70.10', 'K70.11', 'K70.2', 'K70.30', 'K70.31', 'K70.40', 'K70.41', 'K70.9', 'F14.10', 'F14.120', 'F14.121', 'F14.122', 'F14.129', 'F14.13', 'F14.14', 'F14.150', 'F14.151', 'F14.159', 'F14.180', 'F14.181', 'F14.182', 'F14.188', 'F14.19', 'F14.20', 'F14.220', 'F14.221', 'F14.222', 'F14.229', 'F14.23', 'F14.24', 'F14.250', 'F14.251', 'F14.259', 'F14.280', 'F14.281', 'F14.282', 'F14.288', 'F14.29', 'F14.90', 'F14.920', 'F14.921', 'F14.922', 'F14.929', 'F14.93', 'F14.94', 'F14.950', 'F14.951', 'F14.959', 'F14.980', 'F14.981', 'F14.982', 'F14.988', 'F14.99', 'T40.5X1A', 'T40.5X2A', 'T40.5X3A', 'T40.5X4A', 'T40.5X5A', 'F10.11', 'F10.21', 'F14.11', 'F14.21', 'T40.5X2D', 'T51.8X2D', 'T51.92XD', 'T40.5X1D', 'T40.5X2D', 'T40.5X3D', 'T40.5X4D', 'T40.5X5D', 'T40.5X1S', 'T40.5X2S', 'T40.5X3S', 'T40.5X4S', 'T40.5X5S', 'T51.8X2S', 'T51.92XS', 'G31.2', 'K70.41', 'G62.1', 'G72.1', 'O99.314', 'O99.315', 'O99.310', 'O99.311', 'O99.312', 'O99.313', 'R78.0', 'R78.2', 'A36.81', 'B33.24', 'I42.0', 'I42.5', 'I42.7', 'I42.9', 'I43.', 'O90.3', 'A38.1', 'A39.50', 'A39.52', 'B26.82', 'B33.20', 'B33.22', 'B58.81', 'I01.2', 'I09.0', 'I40.0', 'I40.1', 'I40.8', 'I40.9', 'I41.', 'I42.3', 'I42.4', 'I42.8', 'I51.4', 'J10.82', 'J11.82', 'I51.81', 'E74.00', 'E74.01', 'E74.02', 'E74.03', 'E74.04', 'E74.09', 'E88.49', 'E88.81', 'E88.89', 'E88.9', 'O90.3', 'A36.81', 'B33.24', 'I42.7', 'E83.110', 'E83.118', 'E83.19' |

**Table S5. Analytic packages and language used for model development and statistical analysis.**

| Programming Language/Package | Version |
|---|---|
| Python | 3.9.5 |
| TensorFlow | 2.8.0 |
| scikit-learn | 0.24.2 |
| pandas | 1.3.1 |
| numpy | 1.19.5 |
